# Estimates of the severity of COVID-19 disease

**DOI:** 10.1101/2020.03.09.20033357

**Authors:** Robert Verity, Lucy C Okell, Ilaria Dorigatti, Peter Winskill, Charles Whittaker, Natsuko Imai, Gina Cuomo-Dannenburg, Hayley Thompson, Patrick GT Walker, Han Fu, Amy Dighe, Jamie T Griffin, Marc Baguelin, Sangeeta Bhatia, Adhiratha Boonyasiri, Anne Cori, Zulma Cucunubá, Rich FitzJohn, Katy Gaythorpe, Will Green, Arran Hamlet, Wes Hinsley, Daniel Laydon, Gemma Nedjati-Gilani, Steven Riley, Sabine van Elsland, Erik Volz, Haowei Wang, Yuanrong Wang, Xiaoyue Xi, Christl A Donnelly, Azra C Ghani, Neil M Ferguson

## Abstract

**Background:** A range of case fatality ratio (CFR) estimates for COVID-19 have been produced that differ substantially in magnitude.

**Methods:** We used individual-case data from mainland China and cases detected outside mainland China to estimate the time between onset of symptoms and outcome (death or discharge from hospital). We next obtained age-stratified estimates of the CFR by relating the aggregate distribution of cases by dates of onset to the observed cumulative deaths in China, assuming a constant attack rate by age and adjusting for the demography of the population, and age- and location-based under-ascertainment. We additionally estimated the CFR from individual line-list data on 1,334 cases identified outside mainland China. We used data on the PCR prevalence in international residents repatriated from China at the end of January 2020 to obtain age-stratified estimates of the infection fatality ratio (IFR). Using data on age-stratified severity in a subset of 3,665 cases from China, we estimated the proportion of infections that will likely require hospitalisation.

**Findings:** We estimate the mean duration from onset-of-symptoms to death to be 17.8 days (95% credible interval, crI 16.9–19.2 days) and from onset-of-symptoms to hospital discharge to be 22.6 days (95% crI 21.1-24.4 days). We estimate a crude CFR of 3.67% (95% crI 3.56%-3.80%) in cases from mainland China. Adjusting for demography and under-ascertainment of milder cases in Wuhan relative to the rest of China, we obtain a best estimate of the CFR in China of 1.38% (95% crI 1.23%-1.53%) with substantially higher values in older ages. Our estimate of the CFR from international cases stratified by age (under 60 / 60 and above) are consistent with these estimates from China. We obtain an overall IFR estimate for China of 0.66% (0.39%-1.33%), again with an increasing profile with age.

**Interpretation:** These early estimates give an indication of the fatality ratio across the spectrum of COVID-19 disease and demonstrate a strong age-gradient in risk.

## Introduction

As of 3rd March 2020, 90,870 cases and 3,112 deaths of the disease COVID-19 caused by a novel coronavirus had been reported worldwide^1^. To date, the majority of these (80,422 cases and 2,946 deaths) have been reported from mainland China with a geographic focus in the city of Wuhan, Hubei province. However, in recent days the rate of increase in cases has been greatest outside China. At present, substantial outbreaks are occurring in the Republic of Korea (4,812 cases), Iran (1,501 cases) and Italy (2,036 cases). However, geographic expansion of the epidemic continues, with cases now reported from all continents^1^.

Clinical studies conducted on hospitalised cases show that the onset of COVID-19 is associated with symptoms commonly associated with viral pneumonia, most commonly fever, cough/sore throat and myalgia/fatigue^2–6^. The case definition adopted in China and elsewhere includes further stratification of cases as severe (defined as tachypnoea (≧30 breaths/ min) or oxygen saturation ≤93% at rest, or PaO2/FIO2 <300 mmHg) and critical (respiratory failure requiring mechanical ventilation, septic shock or other organ dysfunction/failure that requires intensive care)^7^. According to the WHO/China Joint Mission Report, 80% of laboratory-confirmed cases in China up to 20 February 2020 have mild-to-moderate disease – including both non-pneumonia and pneumonia cases; whilst 13.8% developed severe disease and 6.1% developed to a critical stage requiring intensive care^8^. In a study of clinical progression in 1,099 patients^4^, those at highest risk for severe disease and death included people over the age of 60 years and those with underlying conditions (including hypertension, diabetes, cardiovascular disease, chronic respiratory disease and cancer).

Assessing the severity of COVID-19 is critical to determine both the appropriateness of mitigation strategies and to enable planning for healthcare needs as epidemics unfold. However, crude case fatality ratios (CFRs) obtained from dividing deaths by cases can be misleading^9,10^. Firstly, there can be a period of two to three weeks between a case developing symptoms, subsequently being detected and reported and observing the final clinical outcome. During a growing epidemic the final clinical outcome of most of the reported cases is typically unknown. Simply dividing the cumulative reported number of deaths by the cumulative number of reported cases will therefore underestimate the true CFR early in an epidemic^9–11^. This effect was observed in past epidemics of respiratory pathogens – including SARS^12^ and H1N1^9^ influenza – and as such is widely recognised. Thus, many of the estimates of the CFR that have been obtained to date for COVID-19 correct for this effect^13–16^. Additionally, however, during the exponentially growing phase of an epidemic, the observed time-lags between the onset of symptoms and outcome (recovery or death) are censored and naïve estimates of the observed times from symptoms onset to outcome provide biased estimates of the actual distributions. Ignoring this effect tends to bias the estimated CFR downwards during the early growth phase of an epidemic.

Secondly, surveillance of a newly emerged pathogen is typically biased towards detecting clinically severe cases, particularly at the start of an epidemic when diagnostic capacity is limited (Figure 1). Estimates of the CFR may thus be biased upwards until the extent of clinically milder disease is determined^9^. Data from the epicentre of the outbreak in Wuhan, China have primarily been detected through hospital surveillance and hence are likely to represent moderate or severe illness, with atypical pneumonia and/or acute respiratory distress being used to define suspected cases eligible for testing^7^. In these individuals, clinical outcomes are likely to be more severe, and therefore any estimates of the CFR will be higher. Elsewhere in mainland China and outside, countries and administrative regions alert to the risk of infection being imported via travel initially instituted surveillance for COVID-19 with a broader set of clinical criteria for defining a suspected case. These typically include a combination of symptoms (e.g. cough and fever) combined with recent travel history to the affected region (Wuhan and/or Hubei Province)^2,17^. Such surveillance is therefore likely to detect clinically milder cases but, by initially restricting testing to those with a travel history or link, may have missed other symptomatic cases. More recently, as epidemics have taken off in other countries, cases are now being detected in those with no reported travel links to Wuhan/Hubei province through broader surveillance systems. Some of these cases may represent a milder level of severity – including secondary cases identified via contact-tracing or those identified through sentinel surveillance of influenza-like-illness at primary care^18,19^. In contrast, others will represent the severe end of the disease spectrum with an increasing number identified through hospital surveillance (for example, testing of viral pneumonia) or in a few cases, at post-mortem analysis.

**Figure 1:**
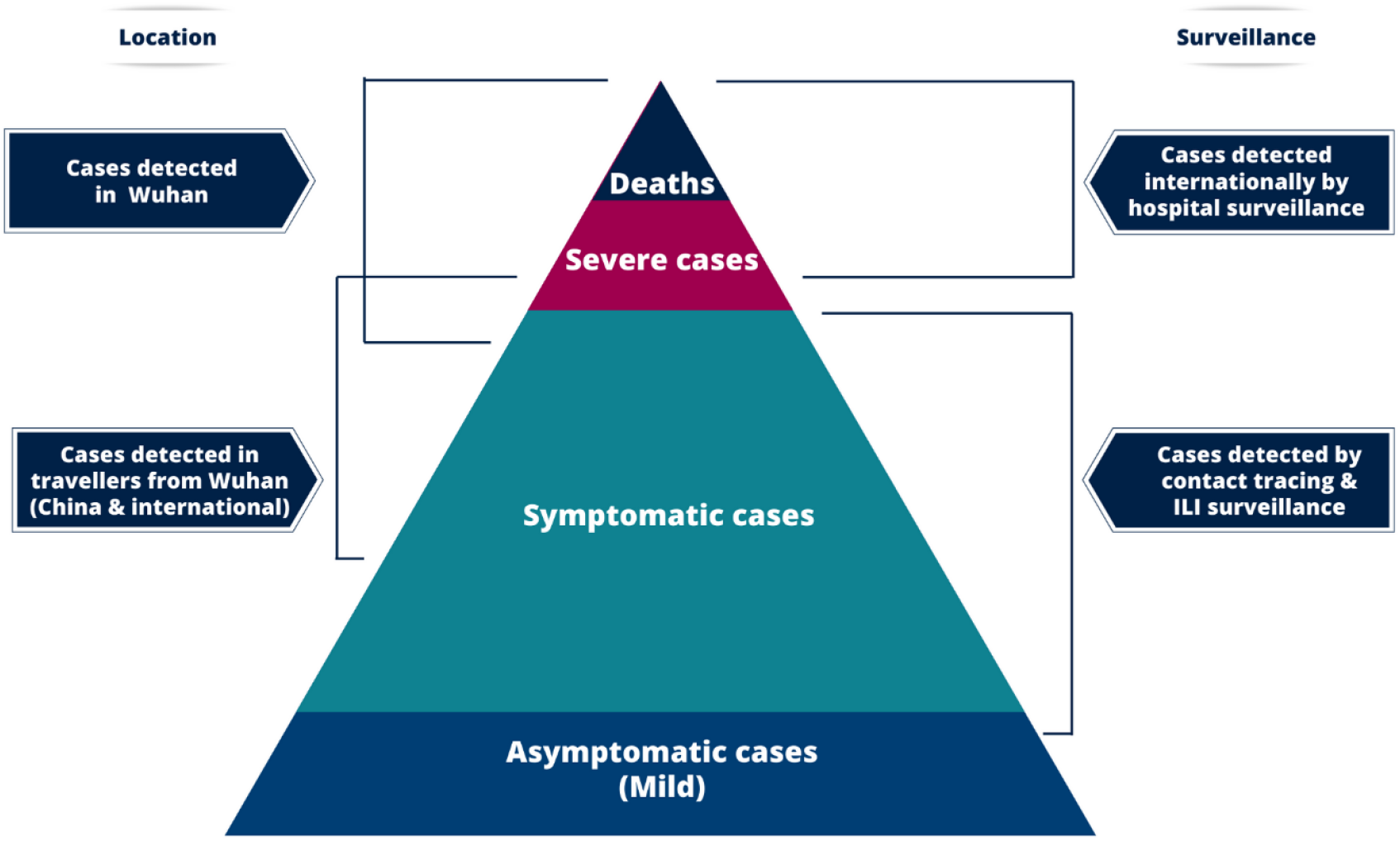
Spectrum of cases for COVID-19. At the top of the pyramid those meeting the WHO case criteria as severe or critical are likely to be identified in the hospital setting presenting with atypical viral pneumonia. These will have been identified in mainland China and amongst those categorised as local transmission internationally. Many more cases are likely to be symptomatic (fever/cough/myalgia) but may not require hospitalisation. These cases will have been identified through links to international travel to high-risk areas and through contact-tracing of contacts of confirmed cases. They may also be identified through population surveillance of, for example, influenza-like illness (ILI). The bottom part of the pyramid represents mild (and possibly asymptomatic) cases. These cases may be identified through contact tracing and subsequently via serological testing.

Quantifying the extent of infection overall in the population (including those infected with either mild, non-specific symptoms or who are asymptomatic, as depicted in the bottom of Figure 1) requires random population surveys of infection prevalence. Serological assays provide the best option for obtaining this denominator; however, robust assays are not currently available. The only such data providing an assessment of the level of infection in a subset of the population not presenting with symptoms at present is therefore the PCR infection prevalence surveys conducted in international residents of Wuhan that have been repatriated to their home country.

Here we attempt to adjust for these biases in data sources to obtain estimates of the CFR (proportion of all cases that will eventually die) and infection fatality ratio (IFR, the proportion of all infections that will eventually die) using both individual-level case report data and aggregate case and death counts from mainland China, administrative regions and international case reports. By adjusting for both underlying demography and potential under-ascertainment at different levels of the severity pyramid (Figure 1), these estimates should be broadly applicable across a range of settings to inform health planning whilst more detailed case data accrue.

## Methods

### Data

#### Individual-level data on early deaths from mainland China

We identified information on the characteristics of 48 cases who died from COVID-19 infection in Hubei Province reported by the National Health Commission and the Hubei Province Health Commission website^20,21^. We recorded the following data elements where available: gender, age, date of symptoms onset, date of hospitalisation, and date of death. Of the 48 cases, neither the date of symptom onset nor the date of report was available for 13 cases. We further removed 8 cases with onset before 01/01/2020 or death before 21/01/2020, and 3 deaths after 28/01/2020 which were the dates consistent with reliable reporting of onset and death in this setting, respectively. This left 24 deaths, which we used to estimate the onset-to-death distribution.

#### Individual-level data on cases outside mainland China

We collated data on 2,010 cases reported from 37 countries and two administrative regions of China (Hong Kong and Macau) from Government (and/or) Ministry of Health websites and media reports up to 25^th^ February 2020. We recorded the following information where available: country or administrative region in which the case was detected, whether the infection was acquired in China or abroad, date of travel, date of symptom onset, date of hospitalisation, date of confirmation, date of recovery, and date of death. We used data from 169 recovered individuals with reported recovery dates and reported/imputed onset dates to estimate the onset to recovery distribution. We used data on 1,334 international cases to obtain estimates of the CFR.

#### Data on aggregate cases and deaths in mainland China

Data on 70,117 PCR-confirmed and clinically diagnosed cases by date of onset in Wuhan and elsewhere in China from 1^st^ January to 11^th^ February 2020 were extracted from the WHO Joint Mission Report^8^. Over this time-period a cumulative total of 1,023 deaths were reported across China, with these data available disaggregated into ten-year age bands between 0-9 and 70-79 years old and those above 80 years old^7^. Using collated data on daily reported deaths obtained each day from the National Health Commission regional websites^21^, we estimate that 74% of deaths occurred in Wuhan and the remainder outside Wuhan. Additionally, the most recent available cumulative estimates (March 3^rd^ 2020) of 80 304 confirmed cases and 2,946 deaths within China were extracted from COVID-19 WHO Situation Report #43^1^.

An earlier pre-print of a subset of these cases up to 26^th^ January 2020 reported the age-distribution of cases categorised by severity for 3,665 cases^22^. Under the China case definition, a severe case is defined as tachypnoea (≧30 breaths/min) or oxygen saturation ≤93% at rest, or PaO2/FIO2 <300 mmHg^7^. Assuming severe cases to require hospitalisation (as opposed to all of the cases that were hospitalised in China, some of which will have been hospitalised to reduce onward transmission), we used the proportion of severe cases by age in these patients to estimate the proportion of cases and infections requiring hospitalisation.

#### Data on infection in repatriated international Wuhan residents

Data on infection prevalence in repatriated expatriates returning to their home countries were obtained from Government (and/or) Ministry of Health websites and media reports. To match to the incidence reported in Wuhan on 30^th^ January 2020, we used data from 6 flights that departed between 30^th^ January 2020 and 1^st^ February 2020 inclusive.

#### Data on cases and deaths on the Diamond Princess cruise ship

In early February 2020 a cruise liner named the Diamond Princess was quarantined after a disembarked passenger tested positive for the virus. Subsequently all 3711 passengers on board were tested over the next month. We extracted data on the ages of passengers onboard on 5^th^ February 2020, the dates of reporting positive tests for 706 PCR-confirmed cases, and date of 7 deaths among these cases from the Japan Ministry of Health, Labour and Welfare reports^23^ and international media.

#### Demography

Age-stratified population data for 2018 were obtained from the National Bureau of Statistics of the People’s Republic of China^24^. According to these data, the population of Wuhan in 2018 was approximately 11 million people.

### Statistical Methods

#### Time intervals between symptom onset and outcome

During a growing epidemic, a higher proportion of the cases will have been infected recently (Figure S1). We accounted for this by re-parameterising a Gamma model to account for exponential growth using a growth rate of 0.14 per day obtained from the early case onset data (see Supplementary Methods). Using Bayesian methods, we fitted a Gamma distributions to data on times from onset-to-death and onset-to-recovery conditional on having observed the final outcome. Missing onset dates were imputed based on dates of report where available.

#### Estimates of the CFR, IFR and proportion hospitalised from aggregated case data

To estimate the CFR using aggregated data we used our estimates of the distribution of times from onset-to-death to project the expected cumulative number of deaths given the onsets observed in Wuhan and outside Wuhan. We began by assuming that the attack rate is uniform across age-groups (i.e. all ages are equally susceptible). Using the age-distribution of the population, for a given attack rate, we therefore obtained an estimate of the expected infections in each age-group. Under-ascertainment was then estimated inside and outside Wuhan by comparing observed cases by age to this expected distribution assuming perfect ascertainment in the 50-59 age-group. For Wuhan, we added an additional scaling (fitted using a Binomial likelihood) to account for further under-ascertainment compared to outside Wuhan (given the over-stretched health system). These steps gave us the expected age-distribution of cases.

For a given onset-to-death distribution, we then obtained a modelled estimate of the cumulative deaths by age under an age-dependent CFR (fitted relative to the CFR in the oldest age-group, which represented the largest number of deaths). This was compared to the observed deaths by age using a Poisson likelihood. These data were then jointly fitted alongside the most-recent age-aggregated cumulative deaths and cases in mainland China, fitted using a binomial likelihood. This follows from the observation that given the current situation, where both observed cases and deaths have dropped substantially following a peak in late January, the ratio of current cumulative cases, once corrected for under-ascertainment, to current death provide a good estimate of the final CFR^11^.

To estimate the IFR we additionally fitted to data on infection prevalence from international Wuhan residents that were repatriated to their home countries. This is formulated as an additional Binomial likelihood, incorporating a translation from incidence to period prevalence accounting for epidemic growth over this period. Our age-stratified CFR and IFR model was then jointly fitted to the case data and infection prevalence data using Bayesian methods. We used our estimates of the onset-to-death distribution obtained from fitting to the 24 deaths in China as a prior distribution. Full mathematical details are given in the Supplementary Information.

The estimates of the proportion of cases that are severe were obtained from a subset of data. Assuming a uniform attack rate by age-groups as before, we use the demography-adjusted under-ascertainment rates calculated above to obtain an estimate of the proportion of infections that require hospitalisation.

To independently validate our IFR estimate, we analysed data from the outbreak on the Diamond Princess cruise liner taking the dates of reported positive tests as a proxy for onset date (acknowledging that this could be before the onset of symptoms for some passengers and after onset for others given potential delays in testing and reporting). We fitted a logistic growth curve to the cumulative proportion testing positive on each day weighted by inverse variance (Supplementary Information) and calculated the expected proportion of deaths observed up to 5^th^ March given the onset times and estimated onset-to-death distribution.

#### Estimates of the case fatality ratio from individual case data

We used parametric (Supplementary Information) and non-parametric methods^11,25^ to estimate the CFR in cases reported outside mainland China using individual-level data on dates of onset, date of report and date of outcome (death, recovery or unknown). Cases where the outcome was unknown were treated as censored observations. For 72% of cases, the date of onset was not reported. For the cases with known date of reporting and missing onset date (n=958) we imputed the date of onset from the observed onset-to-report times. Furthermore, 21% of reports of case recoveries mentioned this in aggregate (e.g. x recoveries on day y) and hence could not be linked to a specific case. This was accounted for in the fitting by including an additional parameter to estimate the proportion of recoveries reported. The parametric models were fitted to the data using Bayesian methods (see Supplementary Material). For the non-parametric method we randomly imputed the missing dates of onset and recovery status 100 times.

All analyses were performed using R software (version 3.6.2) with Bayesian Marko-Chain Monte Carlo (MCMC) fitting using the package drjacoby^26^. Data and code are available on https://github.com/mrc-ide/COVID19_CFR_submission.

## Results

Figure 2A shows the subset of 24 deaths from COVID-19 that occurred in mainland China early in the epidemic. During a growing epidemic, we are more likely to observe shorter times from onset-to-death because those that occur later will not yet have been reported. Correcting for this bias, we estimate the mean time from onset-to-death to be 18.8 days (95% credible interval, crI 15.7-49.7 days) with a coefficient of variation, CV, of 0.45 days (95% crI 0.29-0.54 days). With the limited number of observations in these data and given that they were from early in the epidemic, we could not exclude many deaths occurring with longer times from onset-to-death (hence the high upper credible interval). However, given that the epidemic in China has since declined, our posterior estimate of the mean time from onset-to-death informed by the analysis of aggregated China data is more precise (mean 17.8 days, 95% crI 16.9-19.2 days, Figure 2).

**Figure 2:**
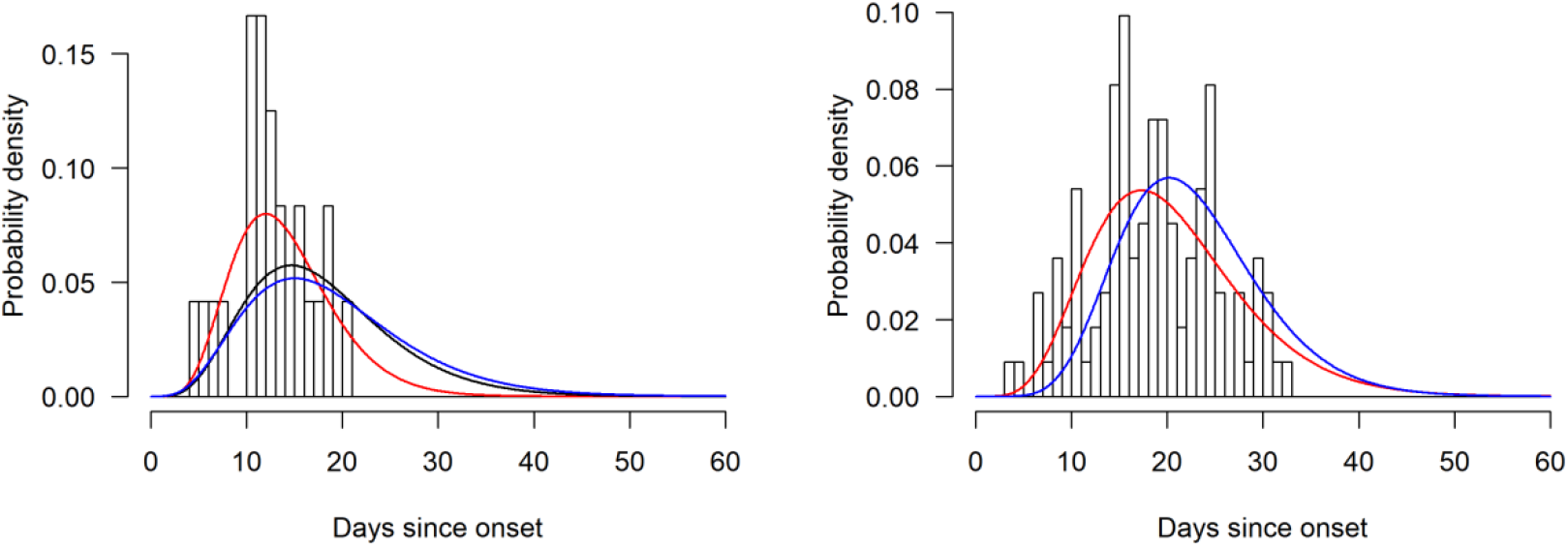
Onset-to-death and onset-to-recovery distributions. (A) Onset-to-death data from 24 cases in mainland China early in the epidemic. (B) Onset-to-recovery data from 169 cases outside mainland China. Red lines show the best fit (posterior mode) Gamma distributions uncorrected for epidemic growth, which are biased towards shorter durations. Blue lines show the same distributions corrected for epidemic growth. The black line on panel (A) shows the posterior estimate of the onset-to-death distribution following fitting to the aggregate case data.

We similarly estimated the mean time from onset-to-recovery using data on outcomes in 169 cases reported outside mainland China. We obtain a slightly longer duration for the onset-to-recovery distribution of 22.6 days (95% crI 21.1-24.4 days) and CV of 0.33 days (95% crI 0.30-0.37 days) (Figure 2).

Table 1 shows estimates of the CFR obtained from aggregate data on cases and deaths in mainland China. A large proportion of the cases, including all of those early in the epidemic, were reported from Wuhan where the local health system was quickly overwhelmed. Furthermore, the case definition outside Wuhan required a travel-link to Wuhan. Figure 3A shows a difference in the resulting age-distribution of cases reported from these two areas. Reported cases in Wuhan are more frequent in older age-groups, perhaps reflecting higher severity (and hence prioritisation for hospitalisation in Wuhan) whilst cases outside Wuhan may also represent a bias in terms of the relationship between age and travel. Adjusting for differences in underlying demography and assuming no overall difference in the attack rate by age, we estimate a high level of under-ascertainment of cases in younger age-groups in both Wuhan and outside Wuhan (Figure 3 C-D). Furthermore, we estimate a higher level of under-ascertainment overall in Wuhan compared to outside Wuhan (Figure 3C). Accounting for this under-ascertainment, we estimate the highest CFR in the 80+ age-group of 13.4% (11.2-15.9%) (Figure 3B, Table 1). Our estimates of CFR decline rapidly with decreasing age, with very low rates in young adults and children.

**Table 1:** Estimates of the case fatality ratio (CFR) and infection fatality ratio (IFR) obtained from aggregate time-series of cases occurring in mainland China. Cases and deaths are aggregate numbers reported from 1^st^ January to 11^th^ February 2020^8^. The crude CFR is calculated as deaths/laboratory-confirmed cases. Our estimates also include clinically diagnosed cases (a scaling of 1.31 applied across all age-groups as the breakdown by age was not reported) which gives a larger denominator and hence lower CFR. The clinically-confirmed cases were not reported by age. The adjusted CFR accounts for the underlying demography in Wuhan/elsewhere in China and corrects for under-ascertainment as in Figure 3. The IFR is obtained by combining estimates of the CFR with information on infection prevalence obtained from those returning home on repatriation flights.

|  | Deaths | Laboratory confirmed cases* | Crude CFR (mean, 95% confidence interval) | CFR adjusting for censoring (posterior mode, 95% credible interval) | CFR additionally adjusted for demography and under-ascertainment (posterior mode, 95% credible interval) | IFR (posterior mode, 95% credible interval) |
| --- | --- | --- | --- | --- | --- | --- |
| Overall | 1023 | 44,672 | 2.29% (2.15,2.43) | 3.67% (3.56,3.80) | 1.38% (1.23,1.53) | 0.657% (0.389,1.33) |
| Age: |  |  |  |  |  |  |
| 0 to 9 | 0 | 416 | 0% (0,0.88%) | 0.095% (0.011,1.34) | 0.0026% (0.0003,0.038) | 0.0016% (0.000185,0.0249) |
| 10 to 19 | 1 | 549 | 0.182% (0.004,1.0) | 0.352% (0.066,1.74) | 0.0148% (0.003,0.076) | 0.007% (0.0015,0.050) |
| 20 to 29 | 7 | 3,619 | 0.193% (0.078,0.40) | 0.296% (0.158,0.662) | 0.06% (0.032,0.132) | 0.031% (0.014,0.092) |
| 30 to 39 | 18 | 7,600 | 0.237% (0.14,0.374) | 0.348% (0.241,0.577) | 0.146% (0.103,0.255) | 0.084% (0.041,0.185) |
| 40 to 49 | 38 | 8,571 | 0.44% (0.31,0.61) | 0.71% (0.52,0.97) | 0.30% (0.22,0.42) | 0.16% (0.076,0.32) |
| 50 to 59 | 130 | 10,008 | 1.3% (1.1,1.5) | 2.1% (1.7,2.4) | 1.3% (1.0,1.6) | 0.60% (0.34,1.3) |
| 60 to 69 | 309 | 8,583 | 3.6% (3.2,4.0) | 5.8% (5.2,6.3) | 4.0% (3.4,4.6) | 1.9% (1.1,3.9) |
| 70 to 79 | 312 | 3,918 | 8.0% (7.1,8.9) | 12.7% (11.5,13.9) | 8.6% (7.5,10.0) | 4.3% (2.5,8.4) |
| 80+ | 208 | 1,408 | 14.8% (13.0,16.7) | 23.3% (20.3,26.7) | 13.4% (11.2,15.9) | 7.8% (3.8,13.3) |
| Age categories: |  |  |  |  |  |  |
| Under 60 | 194 | 30,763 | 0.63% (0.55,0.73) | 1.0% (0.9,1.2) | 0.32% (0.27,0.38) | 0.15% (0.09,0.32) |
| 60 and over | 829 | 13,909 | 5.96% (5.57,6.37) | 9.5% (9.1,10.0) | 6.4% (5.7,7.2) | 3.3% (1.8,6.2) |

**Figure 3.**
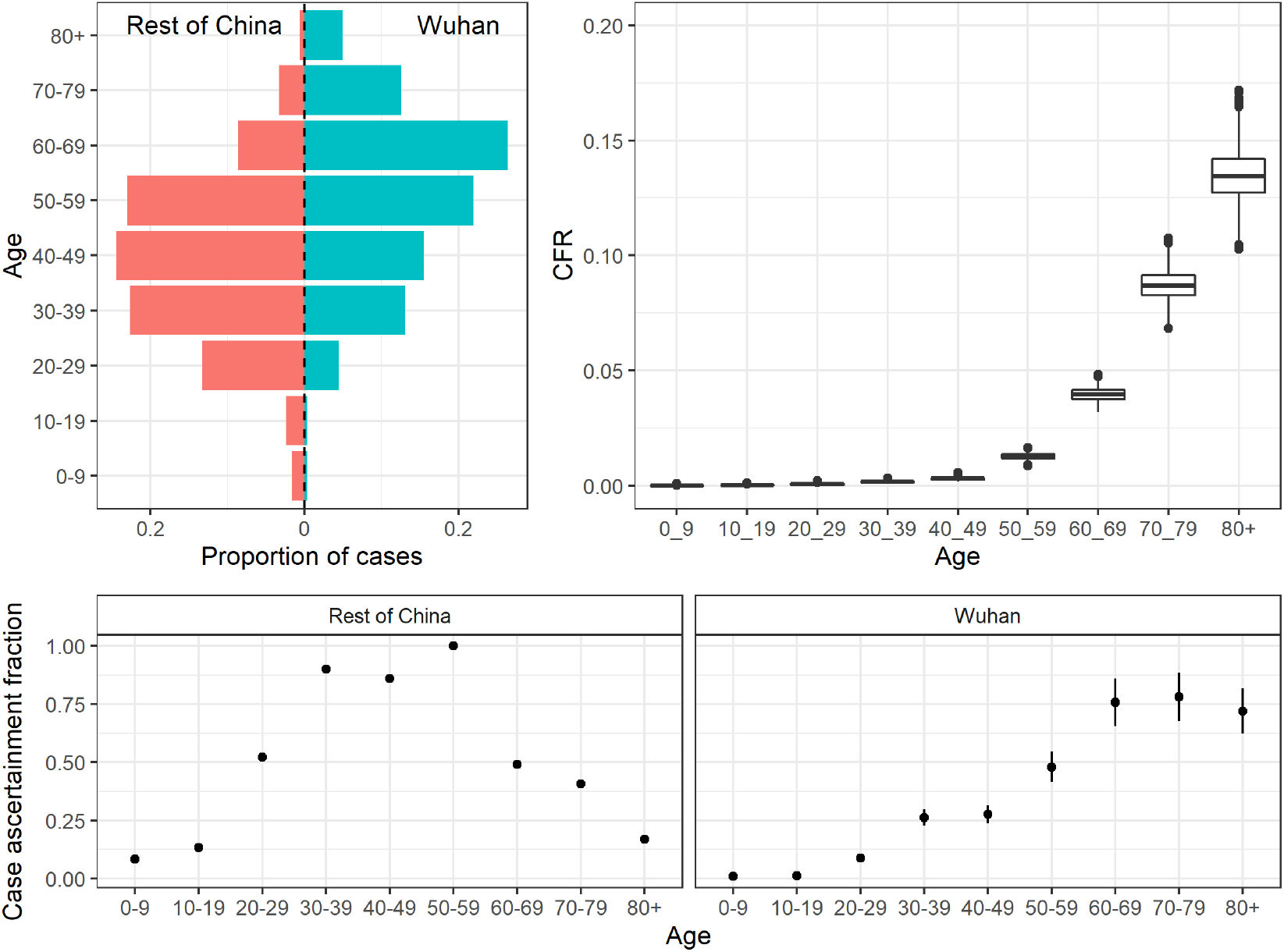
Estimates of the case fatality ratio (CFR) by age obtained from aggregate data from mainland China. A) The age-distribution of cases in Wuhan and elsewhere in China. B) Estimates of the CFR by age-group adjusted for demography and under-ascertainment. C) and D) Estimated proportion of cases ascertained in Rest of China (C) and in Wuhan (D) relative to the 50-59 age-group elsewhere in China.

In cases reported outside mainland China, we estimate an overall modal CFR of 2.7% (95% crI 1.4%-4.7%) using the parametric model (Table 2). However, this reflects a balance of cases detected in different ways in the surveillance system. In those that reported travel to mainland China (and hence will have been detected in the surveillance system due to this travel link), we estimate an overall modal CFR of 1.2% (95% crI 0.4%-4.0%). In those without any reported travel to China (therefore detected either through contact tracing or through hospital surveillance) we estimate a slightly higher CFR (although with high overlap in posterior distributions) of 3.6% (95% crI 1.9%-7.2%) using the parametric model. We estimate a lower CFR in those aged under 60 years of age (1.3%, 95% credible interval, crI 0.5%-3.5%) compared to those aged 60 years and over (4.1%, 95% crI 1.8%-11.0%) in this population. Similar estimates were obtained using non-parametric methods (Table 2). Comparing those under and over-60 years of age, we find estimates that are consistent with those observed in cases from mainland China (Tables 1 and 2).

**Table 2:** Estimates of the case fatality ratio (CFR) obtained from individual-level data on cases identified outside mainland China. The parametric estimates are obtained using the Gamma-distributed estimates of onset-to-death and onset-to-recovery shown in Figure. Non-parametric estimates are obtained using a modified Kaplan-Meier method^11,25^. For both sets of estimates, missing onset dates were multiply imputed using information on the onset-to-report distribution. Note that due to missing data on age and travel status, numbers in the stratified analysis are lower than for the overall analysis. In addition, the parametric method requires a correction for the epidemic growth-rate. The parametric estimates were therefore obtained from the subset of data for which the travel/local transmission and age was known. These samples sizes are shown in brackets.

|  | N | CFR - Parametric | CFR - Non-Parametric |
| --- | --- | --- | --- |
|  |  | Posterior Mode (95% Credible Interval) | Maximum likelihood estimate (95% confidence interval) |
| Overall | 1334 (583) | 2.7% (1.4%-4.7%) | 4.1% (2.1%-7.8%) |
| Travel versus local transmission |  |  |  |
| Travellers to mainland China | 203 | 1.2% (0.4%-4.0%) | 2.4% (0.6%-8.5%) |
| Local transmission | 380 | 3.6% (1.9%-7.2%) | 3.8% (1.7%-8.2%) |
| Age |  |  |  |
| Under 60 | 449 (383) | 1.3% (0.5%-3.5%) | 1.5% (0.6%-3.9%) |
| 60 and over | 181 (158) | 4.1% (1.8%-11.0%) | 12.8% (4.1%-33.5%) |

We estimated a prevalence of infection in international Wuhan residents repatriated on 6 flights of 0.87% (6/689, 95% CI 0.32%-1.9%). Adjusting for demography and under-ascertainment, we estimate an IFR of 0.66% (95% crI 0.39-1.33%). As for the CFR, this is strongly age-dependent with estimates rising steeply from age 50 upwards (Table 1). The demography- and under-ascertainment adjusted proportion of infections requiring hospitalisation ranges from 1.1% in the 20-29 age-group up to 18.4% in the 80+ age-group (Table 3). Using these age-stratified IFR estimates, we estimate the IFR in the Diamond Princess population to be 2.9%. Given the delay from onset of symptoms to death, we would expect 56% of these deaths to have occurred by 5^th^ March 2020 giving an estimate of the current IFR of 1.6% compared to the empirical estimate of 1.0% (7/706, 95% CI 0.4-2.0%).

**Table 3:** Estimates of the proportion of all infections that would be hospitalised obtained from a subset of cases reported in mainland China^22^. We assume that within a UK-context, those that are defined as “severe” would be hospitalised. The rates are adjusted for under-ascertainment and corrected for demography.

| Age | Severe Cases | All Cases | Percentage of infections hospitalised<br>(posterior mode, 95% credible interval) |
| --- | --- | --- | --- |
| 0 to 9 | 0 | 13 | 0% (0.0, 0.0) |
| 10 to 19 | 1 | 50 | 0.04% (0.02, 0.08) |
| 20 to 29 | 49 | 437 | 1.1% (0.62, 2.1) |
| 30 to 39 | 124 | 733 | 3.4% (2.1, 7.0) |
| 40 to 49 | 154 | 743 | 4.3% (2.5, 8.7) |
| 50 to 59 | 222 | 790 | 8.2% (4.9, 16.7) |
| 60 to 69 | 201 | 560 | 11.8% (7.0, 24.0) |
| 70 to 79 | 133 | 263 | 16.6% (9.9, 33.8) |
| 80+ | 51 | 76 | 18.4% (11.0, 37.6) |

## Discussion

Based on extensive analysis of data from different regions of the world, our best estimate at the current time for the CFR of COVID-19 in China is 1.38% (95% crI 1.23%-1.53%). Whilst this remains lower than estimates for other coronaviruses including SARS^27^ and MERS^28^, it is substantially higher than estimates from the 2009 H1N1 influenza pandemic^29–31^. Our estimate of an IFR of 0.66% in China was informed by PCR-testing of international Wuhan residents returning on repatriation flights. This is consistent with the IFR observed to date in passengers on the Princess Diamond Cruise ship. Our estimates of the probability of requiring hospitalisation assume that only severe cases clinically require hospitalisation. This is clearly different from the pattern of hospitalisation that occurred in China, where hospitalisation was also used to ensure case isolation. Mortality can also be expected to vary with the underlying health of specific populations, given that the risks associated with COVID-19 will be heavily influenced by the presence of underlying co-morbidities.

Our estimate of the CFR is substantially lower than the crude CFR obtained from China based on the cases and deaths observed to date, which is currently 3.67%, as well as many of the estimates currently in the literature. The principle reason for this is that the crude estimate does not take into account the severity of cases. Various estimates have been made from patient populations ranging from those with generally milder symptoms (for example international travellers detected through screening of travel history^13,32^) through to those identified in the hospital setting^14,15,33^. We expect higher CFRs in those that are more severely ill. Many case reports note that the presence of other underlying conditions result in poorer prognosis. Although we were unable to adjust for this effect in the absence of detailed individual-level data, this is likely to be correlated with age. However, it should also be noted that the distribution of underlying conditions will vary geographically, and particularly between high-, middle-and low-income settings.

It is clear from the data that has emerged from China that there is a significant increase in the CFR with age. Our results suggest a very low fatality ratio in those under the age of 20. However, as there are very few cases in this age-group, it remains unclear whether this reflects a low risk of death or a difference in susceptibility. Serological testing in this age-group will therefore be critical in the coming weeks to understand the significance of this age-group in driving population transmission. There is a clear increase in the estimated CFR from the age of 50 upwards, with this proportion rising from approximately 1% in the 50-59 age-group to 13% in those aged 80 and above. This increase in severity with age is clearly reflected in case-reports in which the mean age tends to be in the range 50-60 years. Different surveillance systems will pick up a different age-case-mix and we find that those with milder symptoms detected through history of travel are younger on average than those detected through hospital surveillance. Our correction for this surveillance bias therefore allows us to obtain estimates that can be applied to different case-mixes and demographic population structures. However, it should be noted that this correction is applicable under the assumption of a uniform infection attack rate (i.e. exposure) across the population.

Much of the information informing any global estimate of the CFR at the current time is from the early outbreak in Wuhan, China. Given that the health system in this city was quickly over-whelmed, our estimates suggest that there is substantial under-ascertainment of cases in the younger age-groups (who we estimate to have milder disease) in comparison to elsewhere in mainland China. This under-ascertainment is the main factor driving the difference between our estimate of the crude CFR from China (3.67%) and our best estimate of the overall CFR (1.4%). Furthermore, the CFR is likely to be strongly influenced by the availability of healthcare. Whilst in Wuhan this was stretched, our estimates from international cases are of a similar magnitude, suggesting relatively little difference in health outcome. Finally, as clinical knowledge of this new disease accrues, it is possible that outcomes will improve. It will therefore be important to revise these estimates as epidemics unfold.

The world is currently experiencing the early stages of a global pandemic. While China has succeeded in containing spread for two months, this is unlikely to be achievable in most countries. Thus, much of the world will experience very large community epidemics of COVID-19 over the coming weeks and months. Our estimates of the underlying IFR of this infection will inform assessments of health impacts likely to be experienced in different countries and thus decisions around appropriate mitigation policies to be adopted.

## Data Availability

Data and code will be available shortly on GitHub. All data has been sourced from the public domain.

https://github.com/mrc-ide/COVID19_CFR_submission.

## Author Contributions

NMF and ACG conceived the study with input from ID, LCO, RV, PW and CW. ID, LCO and RV led the analysis of individual-case data for the international cases and estimation of the onset-to-outcome distributions with input from CAD, NMF and ACG. RV, PW, CW and PGTW led the analysis of the China data with input from NMF and ACG. NI coordinated management of the team including the data collation and processing. G C-D and NI undertook the extraction of the international case data. HT undertook the extraction of flight repatriation data with input from NI and AD. HF led the extraction of the China mainland data from national and regional websites with input from HW, YW and XX. JTG developed the code for the non-parametric model. ACG produced the first draft of the manuscript. All authors contributed to the final draft.

## Acknowledgements

We are grateful for the input from the following volunteers and hackathon participants from the MRC Centre at Imperial College London: Kylie Ainslie, Sumali Bajaj, Lorenzo Cattarino, Joseph Challenger, Giovanni Charles, Georgina Charnley, Paula Christen, Constance Ciavarella, Victoria Cox, Zulma Cucunubá, Joshua D’Aeth, Tamsin Dewé, Lorna Dunning, Oliver Eales, Keith Fraser, Tini Garske, Lily Geidelberg, Nan Hong, Samuel Horsfield, Min J Kwun, David Jørgensen, Mara Kont, Alice Ledda, Xiang Li, Alessandra Lochen, Tara Mangal, Ruth McCabe, Kevin McRae-McKee, Kate Mitchell, Andria Mousa, Rebecca Nash, Daniela Olivera, Saskia Ricks, Nora Schmit, Ellie Sherrard-Smith, Janetta Skarp, Isaac Stopard, Juliette Unwin, Juan Vesga, Caroline Walters, Lilith Whittles.

## Supplementary Material

### 1 Data Sources

#### 1.1 Temporal Incidence Data for Wuhan and Rest of China

Epidemic curves from Figure 2 of the recent Report of the WHO-China Joint Mission on Coronavirus Disease 2019 (COVID-19) (ref) were digitised and relevant data extracted using the openly available software DataThiefTM: incidence by symptom onset for the period spanning 8th December until 11th February were collated separately for Wuhan and the rest of China. Information on the age-distribution of cases and deaths over the same time period was also extracted from the recent China CDC Weekly Paper (ref) – whilst the deaths were not stratified by location (Wuhan/rest of China), information scraped by volunteers at Imperial College from Chinese provincial Health Commission Reports enabled estimation of the proportion of deaths in China over that time period that had occurred in Wuhan. The observed cases across both locations were then scaled using a number of different adjustments to account for potential underreporting (detailed below). Throughout, we assume all deaths are completely ascertained (i.e. no missed deaths) after the 21^st^ January, and that no detected deaths occurred before that date.

#### 1.2 Individual-Level Data on International Cases

We collated individual line-list data from reports of international cases (see main text). The cases by country are summarised below.

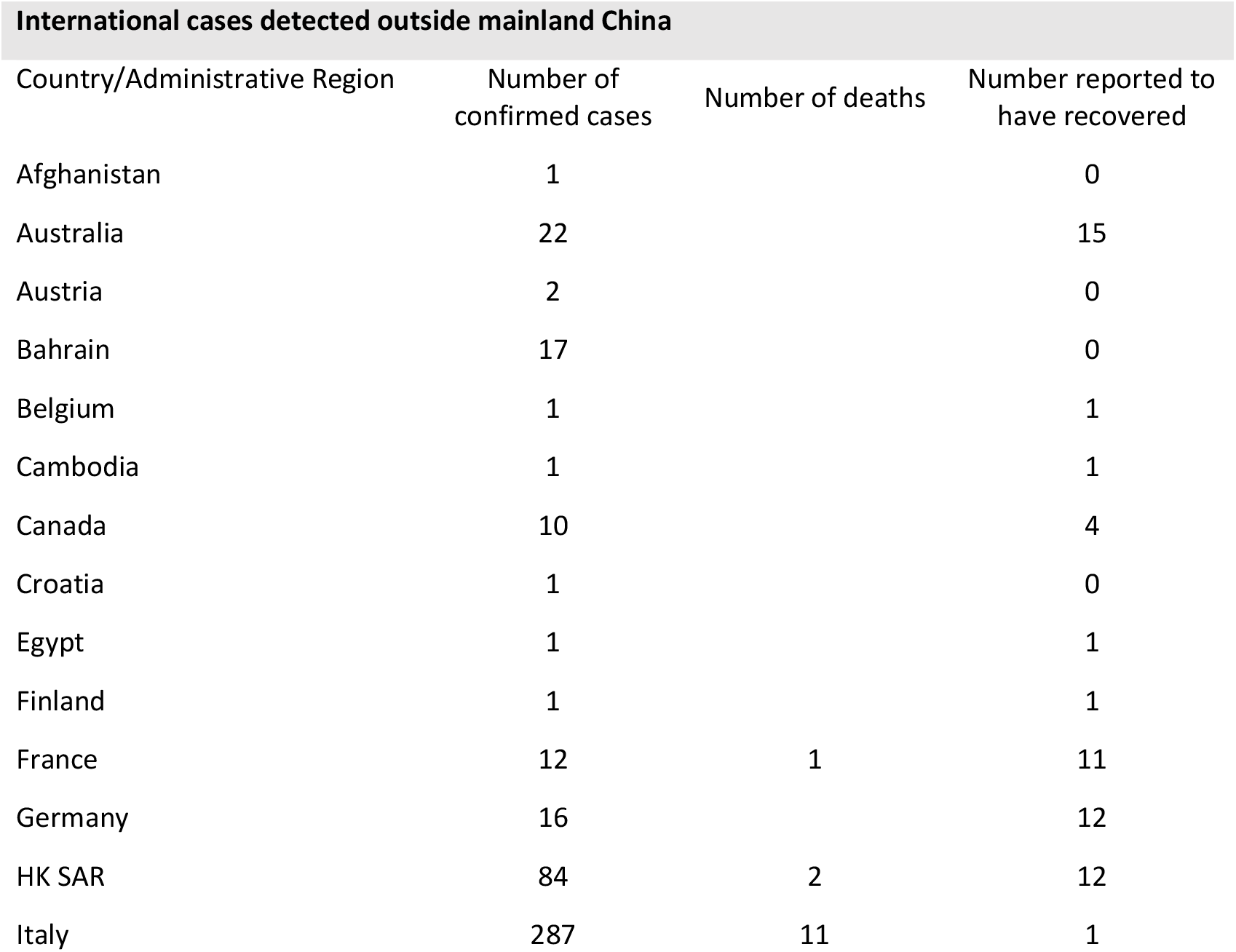
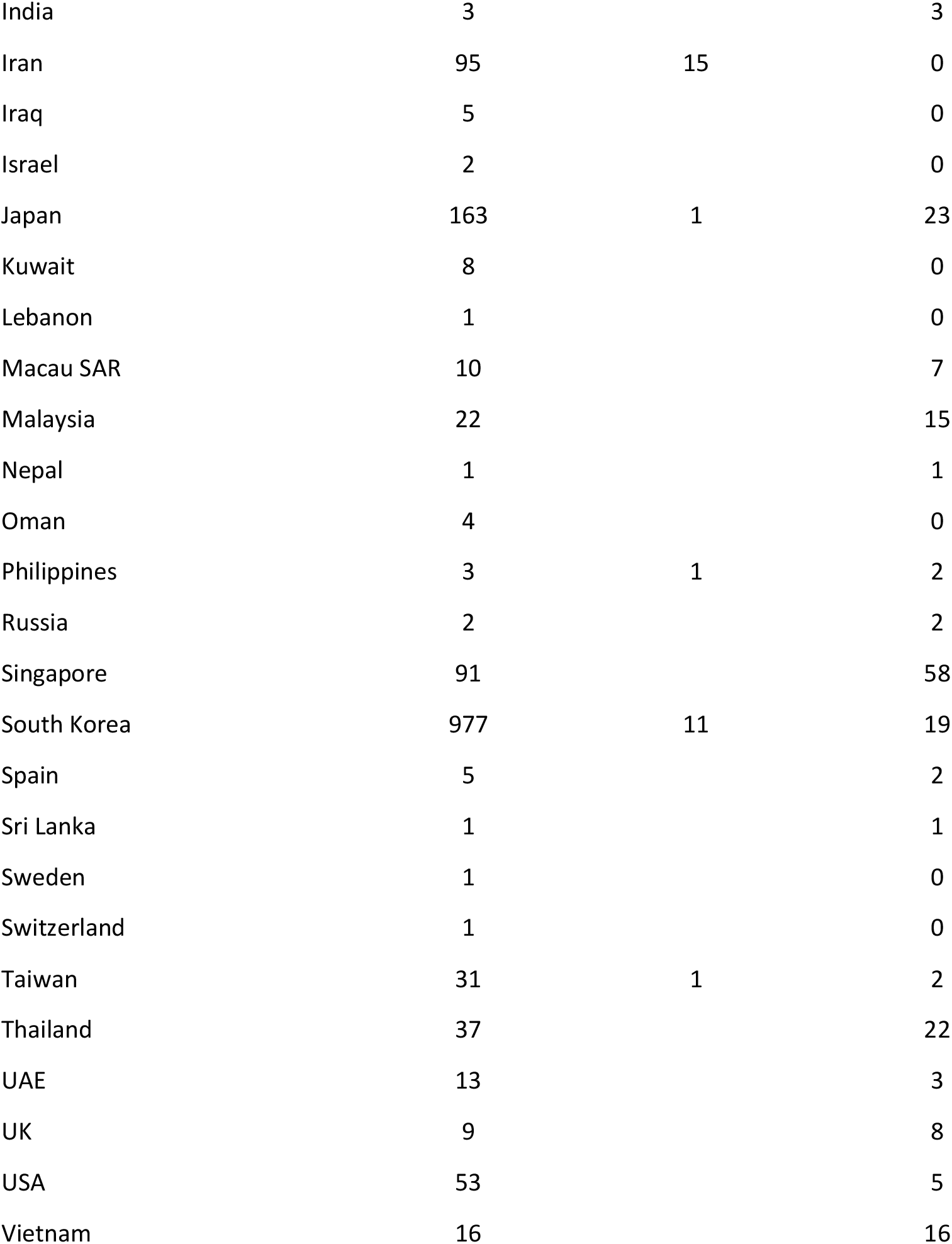

#### 1.3 Prevalence Data from Repatriation Flights

Date on repatriation flights from Wuhan were collated from a number of different sources, including official Ministry of Health reports and media reports. From this data, we considered repatriation flights spanning a three-day period 30^th^ January to 1^st^ February (inclusive) - across these 3 days, a total of 689 individuals were repatriated from Wuhan on flights that tested all individuals (regardless of symptoms) for infection immediately upon arrivals. Testing following this repatriation yielded 6 positive individuals, a point prevalence of 0.87% - this estimate of point prevalence is then incorporated into the analyses detailed below to help estimate the extent of infection underreporting.

| Destination | Date | Number Tested | Number Positive |
| --- | --- | --- | --- |
| Japan | 30/01/2020 | 210 | 2 |
| Japan | 31/01/2020 | 149 | 2 |
| Denmark | 31/01/2020 | 4 | 0 |
| France | 31/01/2020 | 180 | 0 |
| Germany | 01/02/2020 | 115 | 2 |
| Mongolia | 01/02/2020 | 31 | 0 |
| **Total** |  | **689** | **6** |

#### 1.4 Data from Diamond Princess Cruise Ship

We extracted data on the ages of passengers onboard on 5th February, the dates of reporting positive tests for 706 PCR-confirmed cases, and date of 7 deaths. These are shown below.

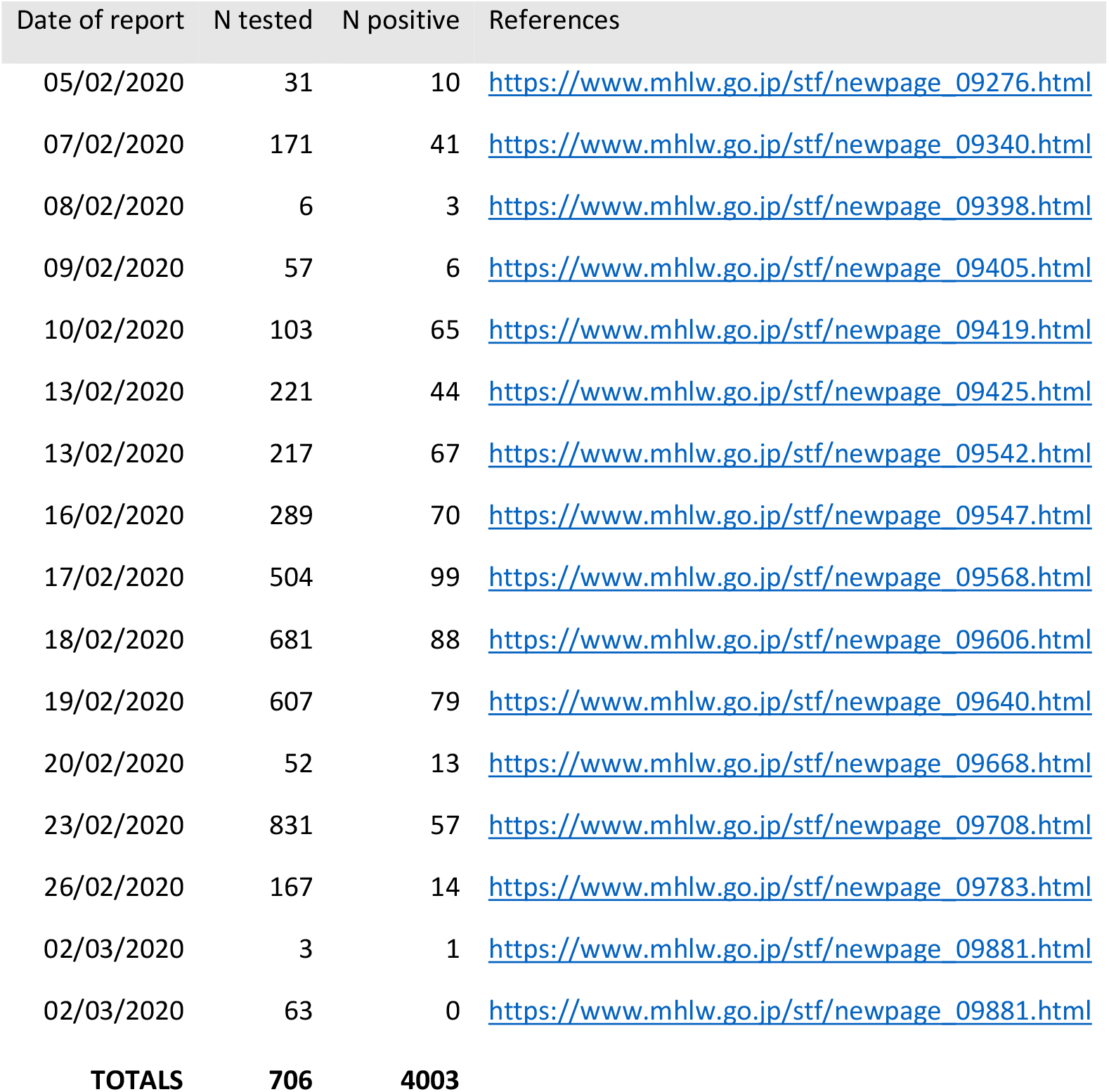

We additionally use the age-distribution of the cases to estimate the IFR. These were available for 531 of the 706 cases; we assumed the age distribution in the remaining cases was the same. These are shown in the table below.

| Age group (years) | Number of passengers | Number testing positive |
| --- | --- | --- |
| 0-9 | 16 | 1 |
| 10-19 | 23 | 2 |
| 20-29 | 347 | 20 |
| 30-39 | 429 | 23 |
| 40-49 | 333 | 25 |
| 50-59 | 398 | 49 |
| 60-69 | 924 | 129 |
| 70-79 | 1015 | 228 |
| 80-89 | 215 | 52 |
| 90-99 | 11 | 2 |
| **Total** | **3711** | **531** |

### 2 Statistical Methods

#### 2.1 Intervals between onset of symptoms and death

Let *t*_*o*_ and *t*_*d*_ be the time (in days) of onset of symptoms and death, respectively, and let *δ*_*od*_ = *t*_*d*_ − *t*_*o*_ be the onset-to-death interval. If *f*_*od*_ (·) denotes the probability density function (PDF) of time from symptom onset to death, then the probability that a death on day *t*_*d*_ had onset of symptoms on day *t*_*o*_ is

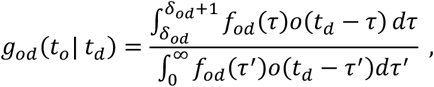

where *o*(*t*) denotes the observed number of onsets that occurred at time *t*. For an exponentially growing epidemic, we assume that *o*(*t*) = *o*_0_ *e*^*rt*^ where *o*_0_ is the initial number of onsets (at *t* = 0) and *r* is the epidemic growth rate. Substituting this, we obtain

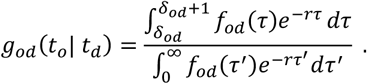

We can use this formula to fit the distribution *g*_*od*_ (·) to the observed data, correcting for the epidemic growth at rate *r* to estimate parameters of the true onset-to-death distribution *f*_*od*_ (·).

If we additionally assume that onsets were poorly observed prior to time *T*_min_ then we can include censoring:

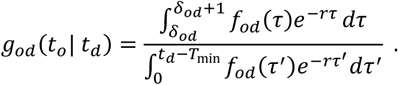

For the special case that we model *f*_*od*_ (·) as a gamma distribution parameterised in terms of its mean *m*_*od*_ and coefficient of variation *s*_*od*_ (defined as the ratio of the standard deviation to the mean), namely *f*_*od*_ (· |*m*_*od*_, *s*_*od*_), it can be shown that

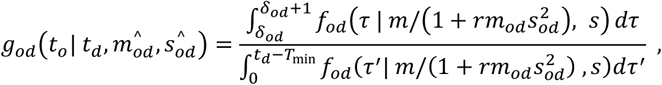

where the transformed mean and standard deviation-to-mean ratios are 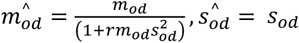.

The Bayesian posterior distribution for *m*_*od*_ and *s*_*od*_ is proportional to the product of this likelihood over a dataset of observed intervals and times of death {*t*_*o*_, *t*_*d*_}:

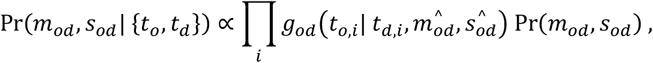

Here Pr(*m*_*od*_, *s*_*od*_) is the joint prior distribution over *m*_*od*_ and *s*_*od*_. We assume a Uniform(10,100) prior on *m*_*od*_ and a Uniform(0.2,0.8) prior on *s*_*od*_, along with a fixed growth rate of *r* = 0.14. We obtained the full posterior distributions of *m*_*od*_ and *s*_*od*_ by computing the joint distribution over a grid in increments of 0.05 and 0.005 respectively. We truncated the distribution by setting the likelihood to zero for combinations of *m*_*od*_ and *s*_*od*_ that generated gamma distributions with 95th percentile >100 days.

#### 2.2 Intervals between onset of symptoms and recovery

Similar to the onset-to-death analysis above, we inferred the onset-to-recovery distribution *f*_*or*_ (· |*m*_*or*_, *s*_*or*_) by fitting to data on the interval *δ*_*or*_ = *t*_*r*_ − *t*_*o*_ between onset of symptoms (*t*_*o*_) and discharge from hospital (*t*_*r*_). As above, we assumed a gamma distribution for *f*_*or*_ (·) resulting in an analytical expression for the epidemic-adjusted distribution *g*_*or*_ (·):

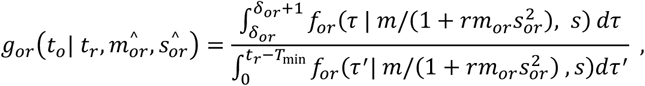

where 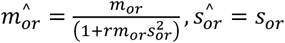.

We used a lower growth rate of *r* = 0.05 for cases in travellers who had been infected in China, where the increase in case numbers had slowed and onsets were earlier. We assumed *r* = 0.14 in locally-acquired cases.

An added complication to this analysis was that many samples had missing onset dates. For samples with missing onset dates we assumed that symptom onset occurred prior to report date, i.e. *t*_*o*_ = *t*_*p*_ − *ε*, where *t*_*p*_ was the date of report (present in all cases) and *ε* was a free parameter. This resulted in an additional set of parameters *ε*_1_, …, *ε*_*n*_, where *n* is the number of cases with missing onset data. Note that when onset data are present, *δ*_*op*_ = *t*_*p*_ − *t*_*o*_ represents observed data, but when onset data are not present this reduces to *δ*_*op*_ = *ε*. Assuming a gamma distribution for the onset-to-report distribution *f*_*op*_ (· | *m*_*op*_, *s*_*op*_) we obtain

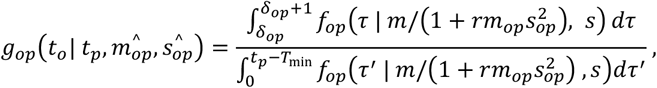

where 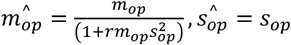.

This likelihoods from the two parts of this analysis were combined and multiplied by the prior to obtain

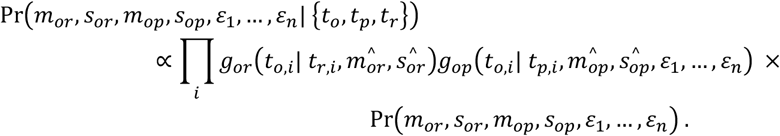

We assumed Uniform(10,100) priors on *m*_*or*_ and *m*_*op*_, and Uniform(0.2,0.8) priors on *s*_*or*_ and *s*_*op*_. We also assumed Uniform(0,50) priors on all *ε* parameters, which were treated as nuisance parameters when summarising other parameters. Due to the high dimensionality of this problem, parameters were estimated via MCMC in the R package drjacoby v1.0^1^.

#### 2.3 Epidemic growth-rate adjustment

The figure below illustrates the requirement for the adjustment for epidemic growth for these onset-to-outcome distributions. We simulated a growing epidemic up to day 60 (number of cases on day 1 = 2, growth rate=0.14, doubling time 5 days). The simulated onset-to-outcome distribution if everyone had been followed up until their outcome was observed is shown by the black bars whilst the onset-to-outcome distribution observed at day 60, censoring those whose outcome is not yet observed is shown by the red bars. The uncorrected Gamma distribution fitted to the observed outcome times at day 60 is shown in red and the Gamma distribution fitted to the observed outcome times at day 60, corrected for epidemic growth, is shown in blue. The latter recovers the true distribution whilst the uncorrected fit results in distribution that is biased towards shorter durations.

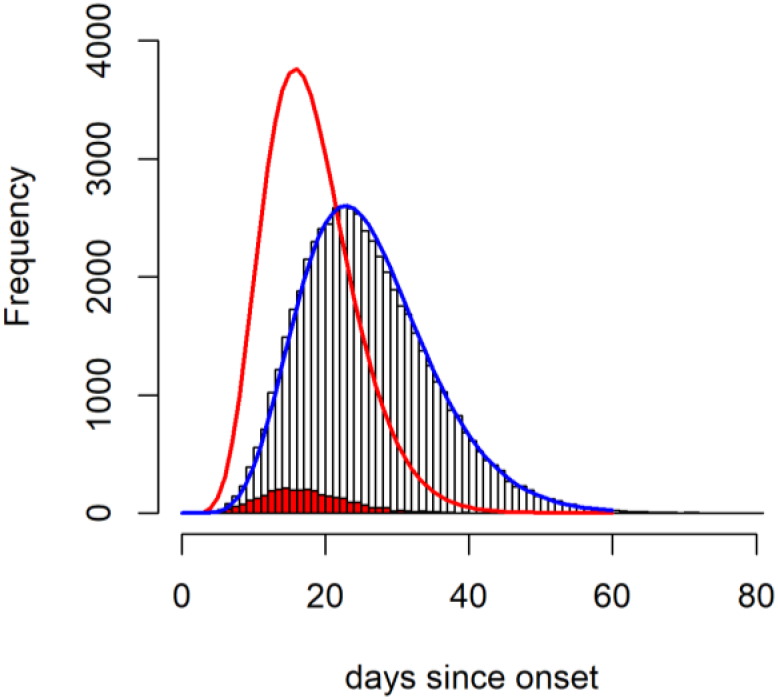

#### 2.4 Age-stratified estimates of the Case Fatality Ratio and Infection Fatality Ratio from aggregate case data

##### 2.4.1 Demographic adjustment

Assuming homogeneous attack rates across the different age groups, the demographic distribution of cases by age across each location should broadly match the demography of the populations in Wuhan and across the rest of China. The reported age-distribution of cases for both locations show striking deviations from the demographic structure of the Chinese populations. Wuhan, in particular, has noticeably fewer cases in younger age groups, and significant overrepresentation of older age-groups (see Figure 1B in main text). Similar patterns are evident in the age-distribution of cases outside China, but to a lesser extent. We hypothesised that these disparities were a product of under-ascertainment of cases, particularly in younger age-groups where a smaller proportion of infections would be expected to be severe and require hospitalisation.

In order to account for these disparities, we adjust the observed cases across both locations (inside Wuhan and outside Wuhan) to produce age-distributions of cases that matches China’s demography. For each age-group and location, we calculate the following:

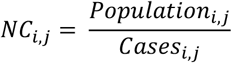

where *i* indexes each age-group and *j* indexes by location, and therefore *Population*_*i,j*_ and *Cases*_*i,j*_ describe the population and number of cases in age-group *i* and location *j* respectively. The reciprocal of *NC*_*i,j*_ is therefore the attack-rate, which describes the number of cases per unit population.

This factor is then used to scale observed cases in the following way. For cases Outside Wuhan, we assume complete ascertainment in the age-group where the attack rate (highest valued reciprocal of

*NC*_*i,j*_) is highest – that of the 50-59 year olds. We then adjust cases in the other age-groups to produce identical attack rates, so that for Outside Wuhan:

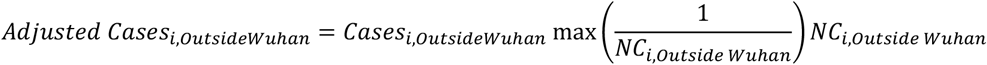

We assume an additional level of under-ascertainment in Wuhan occurring due to the extensive strain on the health system, and so further scale the number of cases after the initial demographic adjustment above, such that

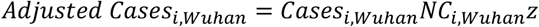

where *z* is a fitted parameter that is smaller than 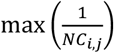, implying that more cases are missed inside Wuhan than in the rest of the country.

We checked the sensitivity of our results to the assumption that there was under-ascertainment outside Wuhan. Under the alternative assumption that cases outside Wuhan are completely ascertained (and hence that the age-distribution observed reflects cases arising only from exposure in Wuhan) we obtained similar estimates (overall CFR 1.87% compared to 1.67% under our baseline assumption).

##### 2.4.2 Statistical modelling framework

We worked within a Bayesian framework in order to jointly estimate the age-stratified case-fatality ratio, the onset-to-death distribution and the true underlying number of cases within Wuhan and other areas of mainland China, incorporating our prior knowledge of the onset-to-death distribution from fitting to observed data from 24 cases from mainland China (see Section 2.1).

Given our case and death age-stratification *A* = {*a* ∈ 1: 9; 1 = 0 − 9 *years old*, 2 = 10 − 19, … 9 = 80 +} we define the following parameters: the associated set of case-fatality rates *θ*_*A*_, mean *m*_*od*_ and standard deviation to mean ratio *s*_*od*_ of the onset-to-death distribution *f*_*od*_ (· | *m*_*od*_, *s*_*od*_). Observed cases are adjusted assuming homogeneous attack rates across age groups and a demographic age-distribution representative of China, assuming perfect case ascertainment in the 50-59 year old age group outside of Wuhan where there were the highest levels of case reporting relative to population size (see above). We also adjust for an additional level of underreporting specific to Wuhan (relative to elsewhere in China), *z*.

To fit these parameters we used the following data: *D*_*w*_, the total observed deaths in Wuhan to 11^th^ February 2020; *D*_*A*_, the total observed deaths by age up to 11^th^ February 2020, including those in Wuhan and *C*_*T,A,L*_, observed cases by day, age and location up to this date. We also incorporated data on the total deaths and cases observed within mainland China by 4^th^ March 2020 (without disaggregation by age or location), *D*_*M*4_ and *C*_*M*4_.

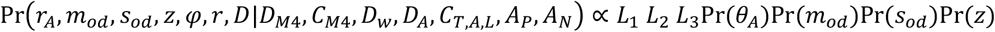

where

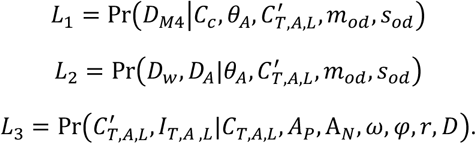

Here term *L*_*1*_ represents the likelihood of the most recently observed crude case-fatality ratio (deaths/cases) in mainland China. The crude case fatality ratio tends to the true case fatality ratio as the proportion of the full epidemic which has been observed increases^2^ and after adjustment for under ascertainment of cases. Cases in China have now reduced substantially relative to their late January 2020 peak. As such, this suggests that the recently estimated crude CFR likely represents a good approximation of the final epidemic CFR. Term *L*_*2*_ represents the likelihood of the observed number of deaths in Wuhan (aggregated across age groups), and also, the observed number of deaths by age across all settings accounting for case-fatality rates by age, the epidemic curve adjusted for differences in ascertainment rates (by age and location) of cases and the distribution between case-onset and death. Term *L*_*3*_ represents the model of how observed cases can be adjusted to reflect true cases, denoted 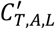, accounting for surveillance capacity in Wuhan, *z*, and age-based disparities in ascertainment throughout the course of the large-scale epidemic.

##### 2.4.3 Estimation of infection rates from flight repatriation data

We also estimate infections, *I*_*T,A, L*_ from true cases accounting for further under-ascertainment present across both locations. We inform this under-ascertainment of all infections using the observed prevalence of infections in travellers (n = 689) repatriated from Wuhan over the time period spanning 30^th^ January – 1st February 2020 (inclusive). We estimate the prevalence of infection in Wuhan on 31^st^ January by:

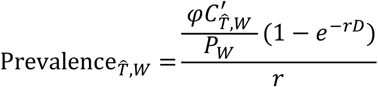

where *φ* is an additional scaling factor for all infections, 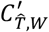 is the estimated incidence of cases on 31^st^ January in Wuhan (after the other age-based and Wuhan specific scalings detailed above), *P*_*W*_ is the population of Wuhan (assumed to be 11,081,000 people), r is the epidemic growth rate (assumed r = 0.14) and D is the detection window (duration that an infection remains detectable). We assume Uniform priors on r of [0,0.1] and D of [7,14].

The remaining terms represent priors which were all uninformative with the exception of the onset-to-death parameters which were set to the likelihood surfaces estimated from the subset of observed onset to death durations.

##### 2.4.4 Capturing age-stratified case-fatality ratios

Setting *T* = 11^th^ of February 2020, the probability a case in age-category *a* with onset date *t* has died by time *T* is:

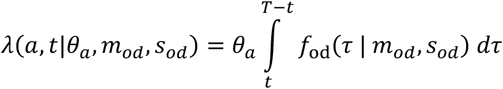

Assuming we observe 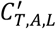, the true number of cases by day and age across all locations from the beginning of the epidemic *t*_0_ = 2^nd^ December (the date our data starts from), the expected number of deaths in age-category *a* is then:

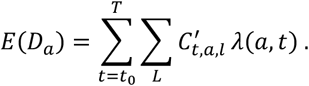

We assume that observed deaths *D*_*a*_ follow a Poisson distribution with rate equal to the expectation *E*(*D*_*a*_):

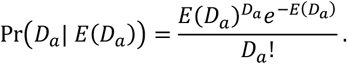

The likelihood of observing the full set of age-specific death-counts observed at *T* is then:

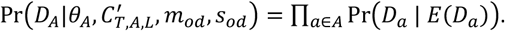

Simultaneously, the expected proportion of cases in Wuhan, *π*_*w*_, can be assumed to follow a Binomial distribution (where *X*∼*Bin*(*N, p*) is the binomial distribution with *X* observations from *N* trials with probability *p*):

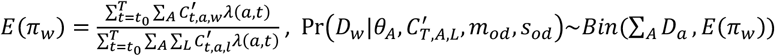

As we assume the age-distribution and location of deaths are independent of one another:

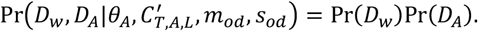

##### 2.4.5 Capturing post-peak overall case-fatality ratio

Given the total number of expected deaths across all-ages according to our age-stratified case-fatality ratios the overall number of expected deaths across all ages in China by *T* is:

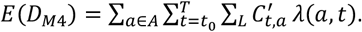

As cases in mainland China have been remained substantially lower than their late January 2020 peak since mid-February, current CFR estimates unadjusted by onset-to-death (i.e. true deaths to date divided by true cases to date) are likely to be a good estimator of the underlying CFR^2^. To capture this information, accounting for our estimates of the underlying surveillance capacity to capture all cases throughout the epidemic, we therefore assume that the current crude CFR in mainland China (i.e. current total deaths as a proportion of the current total observed cases) is a good estimate of the expected deaths arising from cases up to time *T* in China as a proportion of the unadjusted observed cases in this time period:

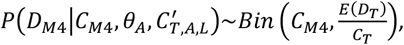

where *C*_*T*_ is the total observed cases in China prior to time *T* (which is 11^th^ February 2020).

#### 2.5 Estimates of the Case Fatality Ratio from individual case data

Continuing our notation from section 2.1, let *t*_*o*_, *t*_*r*_ and *t*_*d*_ denote the times of onset, recovery and death respectively, and let *δ*_*or*_ and *δ*_*od*_ denote onset-to-recovery and onset-to-death intervals. Additionally, let *c* denote the case-fatality ratio (CFR) such that each case has a probability *c* of ultimately resulting in death and a probability (1 − *c*) of ultimately resulting in recovery. We also allow for imperfect identification of recoveries, such that each recovery has a probability *p*_*r*_ of being detected, and a probability (1 − *p*_*r*_) of remaining in the data for an unlimited time as an un-coded or “other” event.

The probability that a patient dies on day *t*_*d*_ given onset at time *t*_*o*_ is given by:

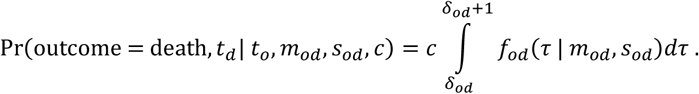

Similarly, the probability that a patient is detected as a recovery on day *t*_*r*_, given onset at time *t*_*o*_, or alternatively recovers but this recovery event is missed, is given by:

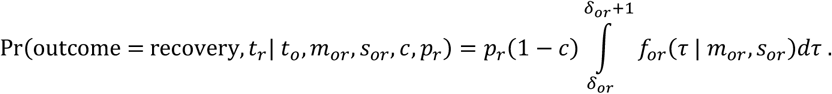

Finally, the probability that a patient remains in hospital at the last date for which data are available, *T*, is

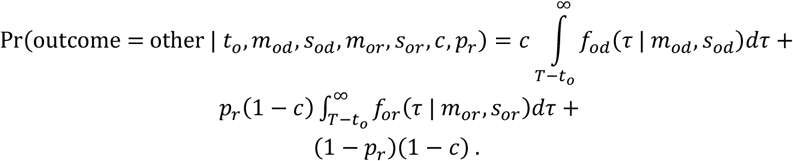

The overall likelihood given all observed outcomes (outcome_*i*_ ∈ {death, recovery, other}) and corresponding outcome times (*t*_*i*_) is simply the product over the individual terms:

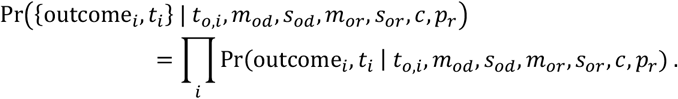

In a Bayesian context, the posterior distribution is obtained by multiplying this likelihood by the priors:

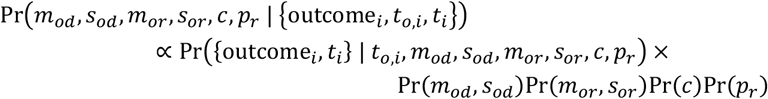

Here we have assumed that the joint prior can be decomposed into separate marginal priors on onset-to-death parameters, onset-to-recovery parameters, and separate priors on *c* and *p*_*r*_. For the first two priors Pr(*m*_*od*_, *s*_*od*_) and Pr(*m*_*or*_, *s*_*or*_) we pass in the posterior distributions from the analyses above, namely the posterior *m*_*od*_ and *s*_*od*_ from the analysis in section 2.1, and the posterior *m*_*or*_ and *s*_*or*_ from the analysis in section 2.4.

#### 2.6 Estimating the infection fatality ratio for the Diamond Princess data

We estimated the proportion of deaths amongst the passengers testing positive on the Diamond Princess that had occurred *π*_*DP*_ (*T*) where *T* was the last date for which data are available (5^th^ March 2020), given the probability density function (PDF) of time from symptom onset to death *f*_*OD*_ (.):

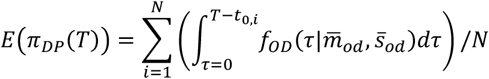

where {*t*_0,*i*_; 1.. *N*} are the set of time of onset in each of the *N* total number of test-positive individuals and 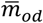 and 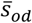 are the posterior mode of the mean and ratio of standard deviation to mean of onset duration distribution obtained from our fitting of these distributions to data from mainland China.

The figure below shows the proportion first testing positive on each date according to Ministry of Health reports, and the fitted logistic growth curve (log odds[proportion positive] = a +b*days, where a=-3.98, b=0.43 and days=days since the first positive test on 5^th^ February up to tests on 2^nd^ March). Initially only symptomatic individuals were tested whilst later testing was extended to all passengers.

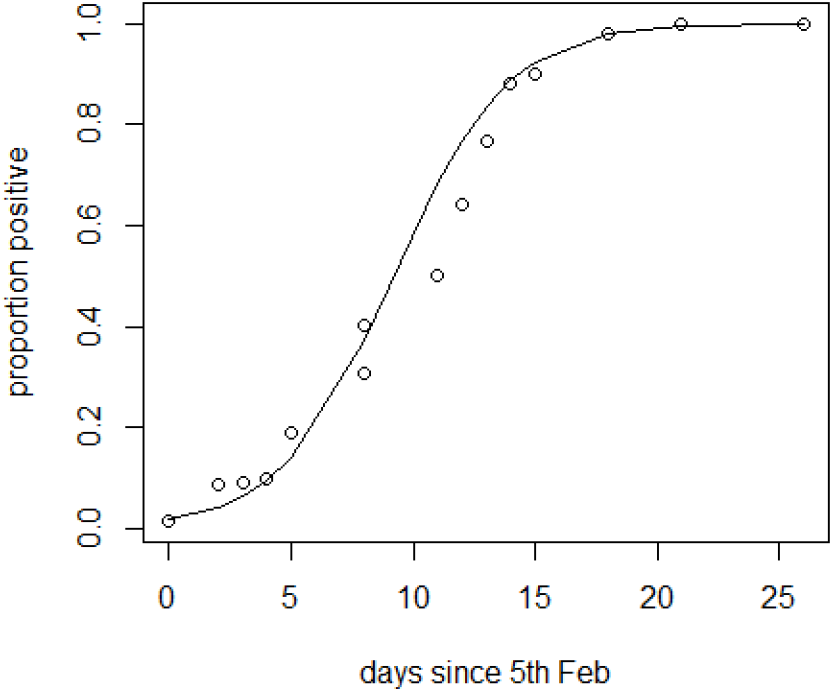

## References

1 World Health Organization. Coronavirus disease 2019 (COVID-19) Situation Report - 43. 2020 https://www.who.int/docs/default-source/coronaviruse/situation-reports/20200303-sitrep-43-covid-19.pdf?sfvrsn=2c21c09c_2.

2 Chan JFW, Yuan S, Kok KH, et al. A familial cluster of pneumonia associated with the 2019 novel coronavirus indicating person-to-person transmission: a study of a family cluster. Lancet 2020; 395: 514–23.

3 Chen N, Zhou M, Dong X, et al. Epidemiological and clinical characteristics of 99 cases of 2019 novel coronavirus pneumonia in Wuhan, China: a descriptive study. Lancet 2020; 395: 507–13.

4 Guan W-J, Ni Z-Y, Hu Y, et al. Clinical Characteristics of Coronavirus Disease 2019 in China. N Engl J Med 2020; : 1–13.

5 Huang C, Wang Y, Li X, et al. Clinical features of patients infected with 2019 novel coronavirus in Wuhan, China. Lancet 2020; published online Jan. DOI:10.1016/s0140-6736(20)30183-5.

6 Li Q, Guan X, Wu P, et al. Early Transmission Dynamics in Wuhan, China, of Novel Coronavirus– Infected Pneumonia. N Engl J Med 2020; published online Jan 29. DOI:10.1056/nejmoa2001316.

7 The Novel Coronavirus Pneumonia Emergency Response Epidemiology Team (China Centre for Disease Control). The epidemiological characteristics of an outbreak of 2019 novel coronavirus diseases (COVID-19) in China. China CDC Wkly 2020; 41: 145–51.

8 World Health Organization. Report of the WHO-China Joint Mission on Coronavirus Disease 2019 (COVID-19). 2020.

9 Garske T, Legrand J, Donnelly CA, et al. Assessing the severity of the novel influenza A/H1N1 pandemic. BMJ 2009; 339: 220–4.

10 Lipsitch M, Donnelly CA, Fraser C, et al. Potential biases in estimating absolute and relative case-fatality risks during outbreaks. PLoS Negl. Trop. Dis. 2015; 9: 1–16.

11 Ghani AC, Donnelly CA, Cox DR, et al. Methods for estimating the case fatality ratio for a novel, emerging infectious disease. Am J Epidemiol 2005; 162: 479–86.

12 Donnelly CA, Ghani AC, Leung GM, et al. Epidemiological determinants of spread of causal agent of severe acute respiratory syndrome in Hong Kong. Lancet 2003; 361. DOI:10.1016/S0140-6736(03)13410-1.

13 Jung S, Akhmetzhanov AR, Hayashi K, et al. Real-Time Estimation of the Risk of Death from Novel Coronavirus (COVID-19) Infection: Inference Using Exported Cases. J Clin Med 2020, Vol 9, Page 523 2020; 9: 523.

14 Mizumoto K, Kagaya K, Chowell G, Yoshida-Nakaadachi-cho U. Early epidemiological assessment of the transmission 1 potential and virulence of 2019 Novel Coronavirus in Affiliations: 5 1 Graduate School of Advanced Integrated Studies in Human Survivability, Kyoto 6. DOI:10.1101/2020.02.12.20022434.

15 Famulare M. 2019-nCoV: preliminary estimates of the confirmed-case-fatality-ratio and infection-fatality-ratio, and initial pandemic risk assessment. 2020.

16 Wu P, Hao X, Lau EHY, et al. Real-time tentative assessment of the epidemiological characteristics of novel coronavirus infections in Wuhan, China, as at 22 January 2020. Euro Surveill 2020; 25: 1–6.

17 Xu X, Wu X, Jiang X, et al. Clinical findings in a group of patients infected with the 2019 novel coronavirus (SARS-Cov-2) outside of Wuhan, China: retrospective case series. Bmj 2020; 368: m606.

18 Jernigan DB, CDC COVID-19 Response Team. Update: Public Health Response to the Coronavirus Disease 2019 Outbreak - United States, February 24, 2020. MMWR Morb Mortal Wkly Rep 2020; 69: 216–9.

19 Bernard Stoecklin S, Rolland P, Silue Y, et al. First cases of coronavirus disease 2019 (COVID-19) in France: surveillance, investigations and control measures, January 2020. Euro Surveill 2020; 25. DOI:10.2807/1560-7917.ES.2020.25.6.2000094.

20 People’s Republic of China. National Health Commission of the PRC, Hubei Province.

21 People’s Republic of China. National Health Commission of the PRC. http://en.nhc.gov.cn/ (accessed Feb 16, 2020).

22 Yang Y, Lu Q, Liu M, et al. Epidemiological and clinical features of the 2019 novel coronavirus outbreak in China. medRxiv 2020; : 2020.02.10.20021675.

23 Government of Japan. Ministry of Health, Labour and Welfare. 2020. https://www.mhlw.go.jp/english/.

24 People’s Republic of China. National Bureau of Statistics China. 2020. http://www.stats.gov.cn/english/Statisticaldata/AnnualData/ (accessed Feb 14, 2020).

25 Griffin J, Ghani A. CASEFAT: Stata module for estimating the case fatality ratio of a new infectious disease. Stat Softw Components 2009; published online May 7.

26 Verity R, Winskill P. drjacoby. https://mrc-ide.github.io/drjacoby/index.html.

27 Lau EHY, Hsiung CA, Cowling BJ, et al. A comparative epidemiologic analysis of SARS in Hong Kong, Beijing and Taiwan. BMC Infect Dis 2010; 10. DOI:10.1186/1471-2334-10-50.

28 Lessler J, Salje H, Van Kerkhove MD, et al. Estimating the severity and subclinical burden of middle east respiratory syndrome coronavirus infection in the Kingdom of Saudi Arabia. Am J Epidemiol 2016; 183: 657–63.

29 Riley S, Kwok KO, Wu KM, et al. Epidemiological characteristics of 2009 (H1N1) pandemic influenza based on paired sera from a longitudinal community cohort study. PLoS Med 2011; 8. DOI:10.1371/journal.pmed.1000442.

30 Kwok KO, Riley S, Perera RAPM, et al. Relative incidence and individual-level severity of seasonal influenza A H3N2 compared with 2009 pandemic H1N1. BMC Infect Dis 2017; 17: 1–12.

31 Wong JY, Wu P, Nishiura H, et al. Infection fatality risk of the pandemic a(H1N1)2009 virus in Hong Kong. Am J Epidemiol 2013; 177: 834–40.

32 Nishiura, Kobayashi Yang, et al. The Rate of Underascertainment of Novel Coronavirus (2019-nCoV) Infection: Estimation Using Japanese Passengers Data on Evacuation Flights. J Clin Med 2020; 9: 419.

33 Wu P, Hao X, Lau EHY, et al. Real-time tentative assessment of the epidemiological characteristics of novel coronavirus infections in Wuhan, China, as at 22 January 2020. Euro Surveill 2020; 25. DOI:10.2807/1560-7917.ES.2020.25.3.2000044.

## References

1 Verity R, Winskill P. drjacoby v1.0. 2020. https://mrc-ide.github.io/drjacoby/index.html.

2 Ghani AC, Donnelly CA, Cox DR, et al. Methods for estimating the case fatality ratio for a novel, emerging infectious disease. Am J Epidemiol 2005; 162: 479–86.

